# First study on surveillance of SARS-CoV-2 RNA in wastewater systems and related environments in Wuhan: Post-lockdown

**DOI:** 10.1101/2020.08.19.20172924

**Authors:** Lu Zhao, Evans Atoni, Yao Du, Huaiyu Zhang, Oscar Donde, Doudou Huang, Shuqi Xiao, Teng Ma, Zhu Shu, Zhiming Yuan, Lei Tong, Han Xia

**Affiliations:** Key Laboratory of Special Pathogens and Biosafety, Wuhan Institute of Virology, Center for Biosafety Mega-Science, Chinese Academy of Sciences, Wuhan 430071, China; University of Chinese Academy of Sciences, Beijing 100049, China; School of Environmental Studies, China University of Geosciences (Wuhan), Wuhan, 430074, China; Central and Southern China Municipal Design & Research Institute Co., Ltd., Wuhan, 430010, China; Department of Environmental Science, Egerton University, P. O. Box 536-20115, Egerton, Kenya

## Abstract

Wastewater-based epidemiology (WBE) has emerged as an effective environmental surveillance tool in monitoring fecal-oral pathogen infections within a community. Congruently, SARS-CoV-2 virus, the etiologic agent of COVID-19, has been demonstrated to infect the gastrointestinal tissues, and be shed in feces. In the present study, SARS-CoV-2 RNA was concentrated from wastewater, sludge, surface water, ground water, and soil samples of municipal and hospital wastewater systems and related environment in Wuhan during the COVID-19 middle and low risk periods, and the viral RNA copies quantified using RT-qPCR. From the findings of this study, during the middle risk period, one influent sample and three secondary treatment effluents collected from Waste Water Treatment Plant 2 (WWTP2), as well as two influent samples from wastewater system of Hospital 2 were SARS-CoV-2 RNA positive. One sludge sample collected from Hospital 4; which was obtained during low risk period, was positive for SARS-CoV-2 RNA. These study findings demonstrate the significance of WBE in continuous surveilling and monitoring of SARS-CoV-2 at the community level, even when the COVID19 prevalence is low. Therefore, the application of WBE is principally useful in tracking the level of infections in communities and the risk assessment of the secondary environment.

## Letter/Short report

Severe Acute Respiratory Syndrome Coronavirus 2 (SARS-CoV-2) is the causative agent for coronavirus disease COVID-19, a current public health crisis of global concern (Zhou et al. 2020). COVID-19 was declared a pandemic by World Health Organization (WHO) on 11^th^ March 2020 (Cucinotta and Vanelli 2020), after its first report in Wuhan to other cities in China and thereafter spread to many other countries. COVID-19 is primarily transmitted via respiratory droplets that people cough, sneeze or exhale, and may also be spread via fomites (Jayaweera et al. 2020; Wang and Du 2020). Moreover, current efforts in the mitigation and prevention of the spread and transmission of SARS-CoV-2 virus has been focused in adoption of non-pharmacological intervention strategies.

In recent times, the assessment of different substances in wastewater has offered vital qualitative or quantitative knowledge on certain populations in a given wastewater catchment, particularly on the drug usage and distribution of drug resistant pathogenic genes within the environment. Moreover, this developing potential application has been propositioned for probable adoption in the field of infectious diseases to track and understand the distribution of disease bio-markers (Choi et al. 2018). Overtime, microbiologists have investigated pathogens in sewerage and wastewater systems, as a public health surveillance tool known as wastewater-based epidemiology (Daughton 2018; Sinclair et al. 2008; Xagoraraki and O’Brien 2020). Wastewater-based epidemiology (WBE) has been applied in surveillance of numerous fecal-oral viruses, foodborne and waterborne pathogens that infected persons typically excrete in high concentration (Bisseux et al. 2018; Iaconelli et al. 2017; Katayama et al. 2008). WBE has been applied in investigating other viruses beyond enteric fecal oral route, since viral shedding involves different body fluids ultimately discharged into the sewage systems (La Rosa et al. 2020).

Therefore, in sight of this global COVID-19 pandemic, scientists from various parts of the world have applied this surveillance system in detecting SARS-CoV-2 RNA particles in wastewater systems (Chavarria-Miró et al. 2020; Haramoto et al. 2020; Kocamemi et al. 2020; Rimoldi et al. 2020; La Rosa et al. 2020). Additionally, in China, a team of scientists from Tongji University investigated the presence of SARS-CoV-2 virus in wastewater discharged from designate hospitals, municipal networks, and downstream in fluent into a sewage plant in Shanghai. In their study findings, SARS-CoV-2 RNA positive samples were detected from wastewater discharges of the designate hospitals (http://www.water8848.com/news/202003/30/123415.html).

Due to the outbreak of SARS-CoV-2 in Wuhan, stringent quarantine measures were enforced throughout the city on January 23, 2020 by the local Hubei government. After 11 weeks of lockdown, traffic control measures begun to be officially lifted on April 8, and life started getting back to normal. As of 7^th^ April, there were 574 confirmed cases and 673 cases under medical surveillance in Wuhan (http://wjw.wuhan.gov.cn/sy/). Indeed, Hubei Centre for Disease Control and Prevention reviewed the emergency response level for COVID-19 in Wuhan, and it was lowered from high to medium risk on 25^th^ March 2020, and thereafter lowered it from medium to low risk on 8^th^ April. Currently there is no published report on tracking of SARS-CoV-2 in wastewater and related environment in Wuhan during the epidemic or after the epidemic.

In the present study, a total of 216 samples that covered middle and low risk periods were collected from various points including wastewater treatment plants, designated hospitals for COVID-19, lake and river that are close to the COVID-19 designated hospitals, between the months of April and May, 2020. Except for the ground water samples, the collection sites were mainly distributed in two districts; with the first one being around Hospital 1-Wastewater treatment plant 1 (WWTP1) in Hongshan district, and the other one being around Hospital 2-Wastewater treatment plant 2 (WWTP2) in Dongxihu district (Figure 1). The samples were transported to the laboratory on ice and processed within 6 h of collection. Raw samples were first centrifuged to remove the debris. Viral particles were concentrated by Polyethylene glycol 8000 (8% w/v, Millipore Sigma) and 0.9 g NaCl (0.3 M, Millipore Sigma), and precipitation done from 40mL supernatant to 1mL. Viral genomes were extracted from 200 μl concentrated samples by using Direct-zol RNA Kit (Zymoresearch), followed by qRT-PCR (NEB) with two primer and probe pairs (RBD2 and ORF1) targeting the RBD and ORF1 genes. Quantification was performed using a standard curve based on the 10-fold dilutions of RNA extracted from SARS-CoV-2 infected VeroE6 culture.

**Figure 1.**
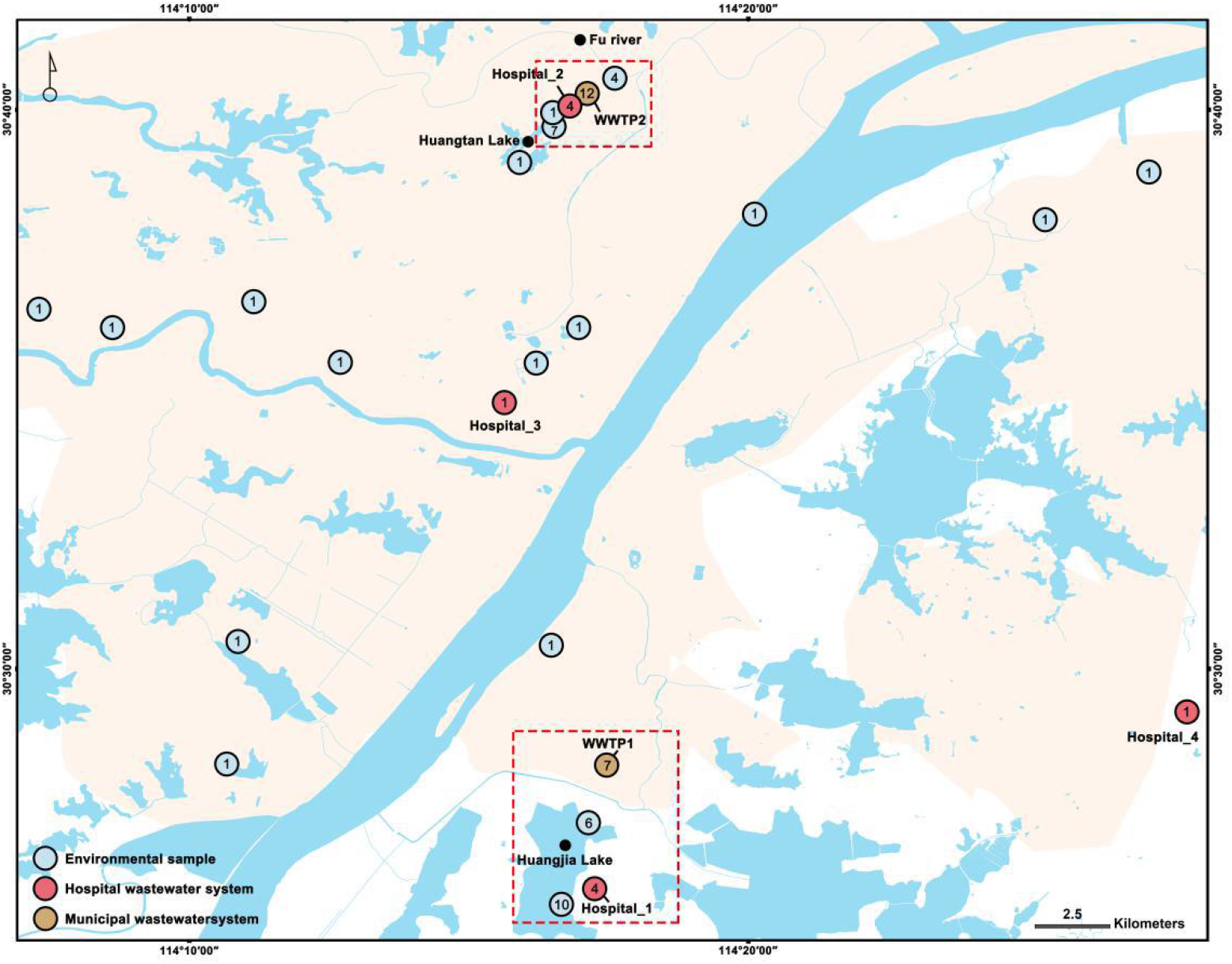
Map of the study area in Wuhan. The number within the circle points stands for the number of sampling sites.

**Figure 2.**
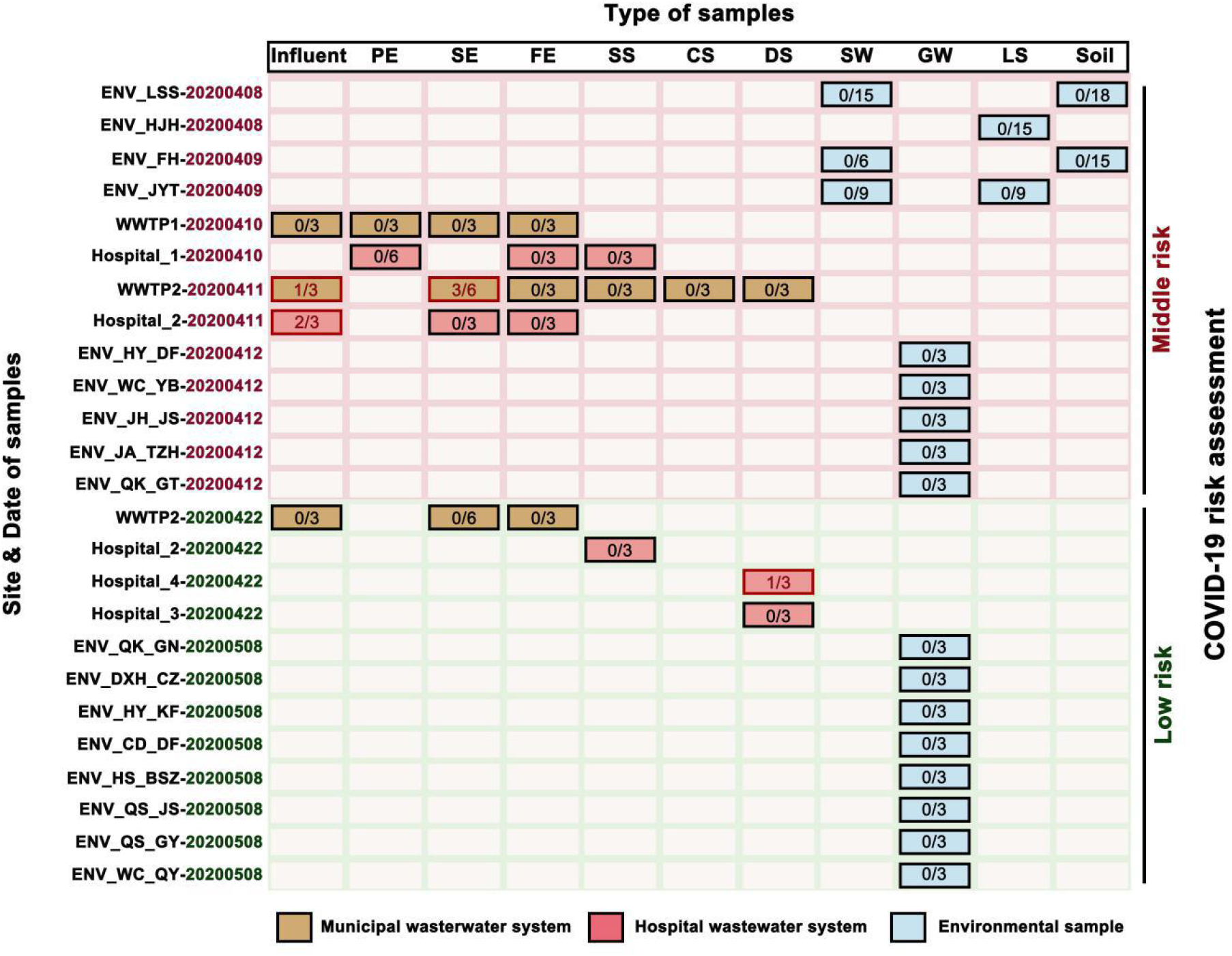
A summary of results for all the tested samples. The numbers in block stand for the number of positive sample over the total number of tested samples.

From the findings of this study, during the middle risk period, positive samples were detected both in municipal and hospital wastewater systems. One of the three influent samples (7.4 ×10^3^ copies/L), three of six secondary treatment effluents (5.3 × 10^3^ and 1.0×10^4^ copies/L) of WWTP2, and two samples from wastewater system influents (3.8 × 10^3^ and 9.3 × 10^3^ copies/L) from Hospital 2 were determined as SARS-CoV-2 RNA positive. During the low risk period, only one positive sludge sample (1.4 × 10^4^ copies/L) was detected in wastewater system handling wastes from Hospital 4 designated for COVID-19 patients. This was consistent with the COVID-19 risk level in Wuhan at the time. Subsequently, attempted viral isolation for all the positive samples was not successful. Further, all of the tested surface and ground waters, lake sediments, and soil samples were SARS-CoV-2 RNA negative in both the middle and low risk periods. This illustrates the positive effect of the stringent epidemiological containment measures that had been implemented in Wuhan, during the high and middle risk phases.

Since the first detection of SARS-CoV-2 in faeces (Zhang et al. 2020), it became clear that human wastewater might contain the novel coronavirus. Hence, WBE of SARS-CoV-2 is crucial because it has been suggested that aerosolization of virus-containing faces might pose a threat in its spread and transmission (Fan Wu et al. 2020). However, whether SARS-CoV-2 is viable under environmental conditions that could facilitate fecal-oral transmission is not yet clear (Lodder and de Roda Husman 2020). Moreover, evidence exists of potential community spread, with the virus spreading easily and sustainably in the community in some affected geographic areas, including China.

Based on recently conducted studies, SARS-CoV-2 RNA has been detected worldwide in influent waters (Ahmed et al. 2020; Lodder and de Roda Husman 2020; Medema et al. 2020; La Rosa et al. 2020; Fuqing Wu et al. 2020), treated wastewater in Paris, France (Wurtzer et al. 2020) and sludge from wastewater treatment Plants (Kocamemi et al. 2020). Therefore, this study findings from Wuhan city are in conformity with these previously conducted studies.

Although SARS-CoV-2 RNA surveillance in wastewaters is a useful WBE drive, the public health risk associated with water cycle is unclear since viral particles infectivity in sewage and faeces is yet to be determined in addition to its probable fecal-oral transmission. Indeed, a recently conducted study has inferred that risk of infection from wastewater and river is insignificant due to the low success rate in cell culture of SARS-CoV-2 from water samples in spite of the high RNA copies (Rimoldi et al. 2020).

Certainly, the findings of this present study offer proof on the sensitivity and probable reliability of environmental surveillance in the detection of emerging disease outbreaks in the population. The sampling was done in late April, when the COVID-19 epidemic was reducing in Wuhan, post-lockdown and massive epidemiological containment measures had been implemented prior. Therefore, the detection of SARS-CoV-2 viral RNA in the tested samples is not surprising because the samples were collected from two hospitals and a wastewater treatment plant, points to the sensitivity of WBE in tracking the pathogen and the need for its adoption in pathogen detection, even under low prevalence record of human illnesses. This phenomenon, in which virus can be detected in sewage in spite of the low prevalence record of human illnesses, might be linked with the capability of wastewater-based surveillance to estimate after thorough epidemiological models, mild, subclinical, or asymptomatic cases (La Rosa et al. 2020). These infected individuals shed viruses into local sewage systems and contribute to virus circulation while remaining substantially undetectable by clinical surveillance, a phenomenon known as the “surveillance pyramid” (Martínez Wassaf et al. 2014). However, one limitation of this study was that data for the high-risk period was lacking. This was because surveillance and detection of SARS-CoV-2 was a prime priority due to the peak in COVID-19 infection in Wuhan. Moreover, there was no regulation that guided WWTP on periodic collection and storage of water samples over a period of time.

In conclusion, the detection of SARS-CoV-2 RNA in various wastewater systems and related environmental samples in Wuhan shows the significance of WBE in continuous surveilling and monitoring SARS-CoV-2 at the community level, under low prevalence record of human illnesses, in contrast to clinical surveillance. This application is principally useful in remote communities and confined populations where mass sampling for the entire population may not be easily achievable at the onset due to inadequate resources or occurrence of asymptomatic patients, though effective sampling techniques is of great essence for achieving accurate results.

## Data Availability

All data is available upon request to the corresponding authors.

## Acknowledgment

We sincerely thank Prof. Hongping Wei and the entire team at the National Biosafety Laboratory in Wuhan, China for the support they extended to us. This work was supported by the Wuhan Bureau of Science and Technology (202002020101010022).

